## Supplemental Table 1 for "Prevalence of, and risk factors for, diabetes and prediabetes in Bangladesh: Evidence from the national survey using a multilevel Poisson regression model with a robust variance"

**Supplementary Table 1: Factors associated with diabetes and prediabetes in adults aged ≥18 years in Bangladeshi population, BDHS 2017-18 (all models)**

| **Characteristics** | **Diabetes, PR (95% CI)** | | | **Pre-diabetes, PR (95% CI)** | | |
| --- | --- | --- | --- | --- | --- | --- |
|  | **Model 2** | **Model 3** | **Model 4** | **Model 2** | **Model 3** | **Model 4** |
| **Individual level** |  |  |  |  |  |  |
| ***Age in years, (ref: 18-34)*** |  |  |  |  |  |  |
| 35-39 | 1.87 (1.51-2.33) | 1.79 (1.44-2.22) | 1.80 (1.45-2.25) | 1.20 (1.02-1.42) | 1.18 (1.00-1.39) | 1.19 (1.01-1.41) |
| 40-44 | 2.44 (1.92-3.09) | 2.33 (1.84-2.93) | 2.37 (1.87-3.00) | 1.22 (1.00-1.48) | 1.20 (0.98-1.45) | 1.23 (1.01-1.48) |
| 45-49 | 2.56 (2.02-3.25) | 2.32 (1.84-2.93) | 2.39 (1.89-3.01) | 1.24 (1.04-1.48) | 1.18 (0.99-1.41) | 1.22 (1.02-1.45) |
| 50-54 | 3.34 (2.59-4.30) | 3.07 (2.39-3.93) | 3.16 (2.45-4.06) | 1.14 (0.90-1.43) | 1.09 (0.87-1.37) | 1.12 (0.90-1.41) |
| 55-59 | 2.95 (2.31-3.77) | 2.59 (2.04-3.29) | 2.70 (2.13-3.43) | 1.30 (1.04-1.62) | 1.21 (0.97-1.51) | 1.26 (1.01-1.58) |
| 60-64 | 3.32 (2.63-4.20) | 2.97 (2.35-3.75) | 3.11 (2.46-3.92) | 1.27 (1.01-1.60) | 1.21 (0.96-1.52) | 1.26 (1.00-1.59) |
| ≥65 | 2.99 (2.32-3.85) | 2.64 (2.06-3.37) | 2.77 (2.16-3.55) | 1.25 (1.01-1.53) | 1.17 (0.95-1.44) | 1.21 (0.99-1.49) |
| ***Sex,*** ***(ref: women)*** | 1.22 (1.06-1.41) | 1.19 (1.03-1.38) | 1.17 (1.01-1.36) | 1.02 (0.90-1.14) | 1.00 (0.89-1.13) | 0.98 (0.87-1.10) |
| ***Body Mass Index (kg/m^2^), (ref: normal weight)*** |  |  |  |  |  |  |
| Underweight (<18.5) | 0.78 (0.63-0.97) | 0.84 (0.68-1.04) | 0.83 (0.67-1.03) | 0.96 (0.82-1.11) | 0.98 (0.84-1.14) | 0.97 (0.83-1.13) |
| Overweight (23.0-27.5) | 1.37 (1.19-1.59) | 1.24 (1.07-1.44) | 1.23 (1.06-1.43) | 1.15 (1.02-1.29) | 1.09 (0.96-1.22) | 1.07 (0.95-1.21) |
| Obese (≥27.5) | 1.79 (1.49-2.15) | 1.47 (1.22-1.77) | 1.45 (1.21-1.75) | 1.39 (1.19-1.62) | 1.24 (1.06-1.45) | 1.23 (1.05-1.44) |
| ***Level of education (ref: higher education)*** |  |  |  |  |  |  |
| No education, preschool | 0.70 (0.55-0.88) | 1.00 (0.79-1.26) | 0.93 (0.74-1.18) | 1.01 (0.83-1.22) | 1.24 (1.01-1.52) | 1.16 (0.95-1.42) |
| Primary | 0.92 (0.75-1.13) | 1.22 (0.99-1.50) | 1.16 (0.95-1.42) | 0.93 (0.79-1.10) | 1.10 (0.93-1.32) | 1.04 (0.88-1.24) |
| Secondary | 0.97 (0.81-1.17) | 1.12 (0.93-1.34) | 1.08 (0.90-1.30) | 0.98 (0.83-1.15) | 1.08 (0.92-1.27) | 1.05 (0.89-1.23) |
| ***Currently working,*** ***(ref: no)*** | 0.73 (0.63-0.84) | *0.79 (0.68-0.92)* | *0.81 (0.69-0.94)* | *0.95 (0.84-1.07)* | 0.99 (0.87-1.11) | 1.01 (0.90-1.14) |
| ***Hypertension, (ref: no)*** | 1.48 (1.31-1.69) | 1.45 (1.28-1.65) | 1.47 (1.30-1.68) | 0.98 (0.87-1.10) | 0.96 (0.84-1.08) | 0.98 (0.87-1.10) |
| **Household level** |  |  |  |  |  |  |
| ***Wealth quintile, (ref: lowest)*** |  |  |  |  |  |  |
| Second |  | 1.06 (0.81-1.39) | 1.04 (0.79-1.35) |  | 0.85 (0.69-1.05) | 0.82 (0.66-1.03) |
| Middle |  | 1.31 (1.02-1.69) | 1.24 (0.96-1.60) |  | 0.98 (0.80-1.20) | 0.94 (0.77-1.15) |
| Fourth |  | 1.78 (1.38-2.30) | 1.60 (1.23-2.09) |  | 1.27 (1.05-1.55) | 1.16 (0.95-1.42) |
| Highest |  | 2.56 (1.99-3.26) | 2.21 (1.70-2.86) |  | 1.58 (1.29-1.94) | 1.36 (1.10-1.68) |
| **Community level** |  |  |  |  |  |  |
| ***Place of residence,*** ***(ref: rural)*** |  |  | 1.02 (0.89-1.18) |  |  | 1.00 (0.88-1.14) |
| ***Administrative division, (ref: Barishal)*** |  |  |  |  |  |  |
| Chattogram |  |  | 0.98 (0.74-1.29) |  |  | 0.82 (0.66-1.03) |
| Dhaka |  |  | 1.32 (1.02-1.71) |  |  | 1.19 (0.94-1.49) |
| Khulna |  |  | 0.78 (0.60-1.01) |  |  | 0.59 (0.46-0.77) |
| Mymensingh |  |  | 0.94 (0.70-1.27) |  |  | 0.74 (0.59-0.93) |
| Rajshahi |  |  | 0.90 (0.68-1.20) |  |  | 0.57 (0.43-0.75) |
| Rangpur |  |  | 0.67 (0.50-0.91) |  |  | 0.54 (0.42-0.71) |
| Sylhet |  |  | 1.00 (0.72-1.38) |  |  | 0.73 (0.56-0.94) |
